# Knowledge, attitudes, and perceptions of long-acting reversible contraceptive (LARC) methods among healthcare workers in sub-Saharan Africa: a systematic review and meta-analysis

**DOI:** 10.1101/2020.10.27.20220434

**Authors:** Laura Rouncivell, Simbarashe Takuva, Neo Ledibane, Alfred Musekiwa, Trudy D Leong

**Author notes:** <u>Corresponding Author:</u> Laura Rouncivell. School of Health Systems and Public Health, Faculty of Health Sciences, University of Pretoria, Pretoria, South Africa.

## Abstract

**Objective:** To assess the knowledge, attitudes, and perceptions (KAP) of long-acting reversible contraceptive (LARC) methods among healthcare workers (HCWs) in sub-Saharan Africa (SSA).

**Methods:** A systematic review and meta-analysis were conducted following the PRISMA methodology. Two authors independently searched three electronic databases for studies published between 2000 and January 2020 reporting on the KAP of LARC methods among HCWs in SSA. Titles and abstracts were screened against eligibility criteria, data were extracted and the included studies were assessed for risk of bias. A meta-analysis of proportions for 11 pre-determined questions relating to LARC KAP was performed. Heterogeneity was explored using the I^2^-statistic and publication bias investigated using funnel plots and Egger’s tests.

**Results:** Twenty-two studies comprising of 11 272 HCWs from 11 SSA countries were included. Forty-one percent (95% CI: 20%, 61%) of HCWs had received intrauterine contraceptive device (IUCD) insertion training while 63% (95% CI: 44%, 81%) expressed a desire for training. Only 27% (95% CI: 18%, 36%) deemed IUCD appropriate for HIV-infected women. Restrictions for IUCD and injectables based on a minimum age were imposed by 56% (95% CI: 33%, 78%) and 60% (95% CI: 36%, 84%), respectively. Minimum parity restrictions were observed among 29% (95% CI: 9%, 50%) of HCWs for IUCDs and 36% (95% CI: 16%, 56%) for injectable contraceptives. Heterogeneity was high and publication bias was present in two of the 11 questions.

**Conclusion:** The systematic review and meta-analysis indicate that unnecessary provider-imposed restrictions may hinder the uptake of LARC methods by women in SSA.

**Conflicts of Interest:** None.

**Ethics approval:** Ethical approval was received from the Faculty of Health Sciences Research Ethics Committee (REC) at the University of Pretoria, School of Health Systems and Public Health. Reference Number: 640/2019

**Authors Contributions:** All authors contributed to the design of the study and the preparation of the manuscript. LR, ST and AM contributed toward the statistical analysis. All authors read and approved the content of the manuscript.

## BACKGROUND

The use of and access to family planning and contraception play a critical role in reducing maternal and infant mortality, improving sexual and reproductive health (SRH) outcomes and fulfilling women’s human rights across the globe (1). An important outcome of family planning is the prevention of unintended pregnancies. Despite significant achievements driven by the Family Planning 2020 initiative which saw 121 million unintended pregnancies prevented between July 2019 and July 2020 among the 69 target countries (2), 74 million still occur each year in low and middle-income countries (LMICs) (3). Due to the resulting adverse maternal and foetal health, social and economic consequences, unintended pregnancies pose a significant threat to global health (4). In LMICs, unintended pregnancies reduce the likelihood of women attending the optimal number of antenatal care visits (5) while also predisposing them to unsafe abortions, particularly where access to abortion services is limited (6). Up to 25 million unsafe abortions and 47 000 maternal deaths are attributed each year to unintended pregnancies (3). Recent modelling suggests that the global unintended pregnancy rate for women aged 15-49 years between 2015 and 2019 was 64 per 100 000 with sub-Saharan Africa (SSA) experiencing a rate of 91 per 100 000 (6). Overall, 37% of unintended pregnancies in SSA ended in abortion, including 50% in countries where abortion is completely restricted (6). Sexual and reproductive health policies across SSA are gaining notable attention, aiming to provide universal access to reproductive health services (7). However, a key driver of unintended pregnancies remains the unmet need for family planning, estimated globally in 2019 at 10% and increasing to an average of 17.1% in SSA with 15 countries having an unmet need greater than 20% (8). The resulting effect of a high unmet need for effective family planning propels the rate of unintended pregnancies. Across 36 LMICs between 2005 and 2014, 65% of unintended pregnancies were among women not using contraception or using a traditional method (withdrawal or calendar-based), 31% occurred among women using short-acting reversible contraceptive (SARC) methods and 2.6% among those using a long-acting reversible contraceptive (LARC) method (3).

Long-acting reversible contraceptives are the preferred intervention, as they are user-independent and last for 3-5 years (progestin injectables are also considered to be long-acting, though they last for 2-3 months)(9). Advantages of LARCs over short-acting reversible contraceptives (SARCs) include increased effectiveness (>99%) as seen by their low failure rates (10). LARCs have shown to be highly effective and safe in the prevention of rapid repeat pregnancies (RRP) while the Copper intrauterine device (Cu-IUD) is also indicated as a method of emergency contraceptive. Therefore, these methods play a critical role in the prevention of unsafe abortions and their associated risks (11,12). Despite the pronounced benefits of modern contraception (LARCs or SARCs), the trend in uptake of LARCs in SSA is significantly lower in comparison to other regions of the world (7). In 2015, 25.1% of married women (15-49 years) in Africa were using a modern method of contraception (7). While injectable progestin contraceptives (IPC) account for 45% of modern methods utilised, the uptake of intrauterine contraceptive devices (IUCDs) and sub-dermal implants which yield higher adherence and lower discontinuation rates remain low (13,14). An analysis of eight SSA countries (Chad, Cameroon, Ghana, Malawi, Mali, Rwanda, Zambia and Zimbabwe) indicated that by 2016, only 2.61% of women were using either an IUCD or an implant (15). Total fertility rates (TFR) in the region are also high, including Kenya, Nigeria and the Republic of Niger at 4.8, 5.2 and 7.6 children per woman, respectively (16); highlighting the impact of low LARC uptake. In South Africa, however, the TFR is lower at 2.33 children per woman (17).

The high HIV burden in the SSA population makes effective contraceptive use crucial. Unintended pregnancies and RRP among women living with HIV (WLWHIV) or using antiretroviral therapy (ART) for pre-exposure prophylaxis (PrEP) can result in adverse maternal and infant health consequences or an increase in vertical HIV transmission (18). A recent prospective study in South Africa showed that women with an unintended pregnancy had almost triple the odds of a higher viral load at 36 to 60 months postpartum compared to those with a planned pregnancy (19). This study suggests that the long-term impact of unintended pregnancy among this already vulnerable population can have a significant effect on ART adherence. Additionally, the concern over ART drug interactions with hormonal contraception has also increased (20). According to an updated systematic review, there is evidence indicating reduced efficacy of progestin-based contraceptive implants in conjunction with efavirenz-based regimens. Among these women, pregnancy rates were higher in comparison to those not using efavirenz-based regimens (20). For this reason, the low utilisation of the non-hormonal Cu-IUD in regions with a double burden of HIV and unintended pregnancies is concerning.

The knowledge, attitudes and perceptions (KAP) held by HCWs can greatly impact on quality of care, as highlighted in a systematic review focusing on maternal healthcare providers showing how negative attitudes undermine health-seeking behaviour and adversely impact patient well-being (21). Healthcare workers are women’s first point of contact when seeking family planning services. Therefore, the KAP held by HCWs must align with evidence-based practice to ensure all women can make an informed decision and receive the most effective contraception of their choice. However, from the growing body of evidence exploring HCWs influence on contraception uptake, misconceptions, particularly around the suitability of LARCs for specific female populations including adolescents, WLWHIV and nulliparous women is seen to exist (22–25). With a population surge in SSA predicted by 2050 (26) and current low LARC method uptake, there is a need for SSA to renew its strategy to provide access to contraceptives, in particular, LARCs which can prevent avoidable adverse health outcomes arising from unintended pregnancies. This systematic review and meta-analysis aimed to determine the current state of knowledge, attitudes, and perceptions among HCWs in SSA regarding LARC methods.

## METHODS

A systematic review and proportion meta-analysis (27) were conducted to explore the KAP of LARC methods among HCWs across SSA. The study followed the Preferred Reporting Items for Systematic Reviews and Meta-analyses (PRISMA) guidelines (28). The protocol for this systematic review is registered on the PROSPERO database (ID: CRD42020148302).

### Eligibility criteria

All cross-sectional, longitudinal, population-based and mixed-method studies that investigated all or a combination of the constructs of knowledge, attitudes, and/or perceptions of HCWs towards LARC methods were eligible for inclusion. All studies published in English between January 2000 and January 2020 were considered. Studies were included if they focussed on IUCD contraception, hormonal sub-dermal implants or hormonal injectable contraceptives and assessed the KAP of these by HCW using structured questionnaires. Healthcare workers were defined as medical practitioners, nurses, pharmacists, community health (extension) workers (CHEWs), clinical associates, and family planning providers, who were practising in SSA at the time of the study. Knowledge of LARC methods referred to HCWs understanding of how the methods worked, their training experience, their capacity to counsel clients and their ability to provide the contraceptives. Attitude was defined as the impact of the HCWs own religious, moral and ethical beliefs on their willingness to provide LARC methods and perceptions were considered as the restrictions imposed on the provision of LARCs (e.g. restricting access based on a client’s age or parity, HIV-status or marital status).

### Information sources and search strategy

Two researchers (LR and ST) independently searched three major online databases (PubMed, Ovid (Medline), and Scopus) for studies that met the inclusion criteria. A final search strategy was developed and applied to the databases in October 2019 and again in January 2020. The search strategy (attached in supplementary file 1) utilised Boolean operators (“AND” or “OR”) to ensure the detection of relevant studies. Grey literature was also searched to identify any relevant organisational or governmental reports.

### Study selection

After applying the search strategy, two researchers (LR and ST) independently screened all titles and abstracts that were produced. Of the records that were deemed preliminarily relevant, the full-text articles were obtained and further screening was conducted. The screening of studies was done using the systematic review reference manager app, Rayyan QCRI (29). The relevance of the studies was assessed based on the initial inclusion criteria that were developed. All studies not meeting the criteria were subsequently excluded, with reasons therefore documented. Any discrepancies were discussed and resolved through consensus.

### Data collection and data extraction

A data extraction tool was developed for this study. The tool was piloted on three studies and adjusted where necessary. The tool documented data on study title, year and author, study design, study population, outcome measured, contraceptive focus, method of recruitment, data collection period, sampling technique, response rate, results, and study strengths and limitations. For each study included in the review, a data collection form was completed.

Following extraction, the data were reviewed to identify common outcomes among the studies. From this, 11 questions (table 1) were developed by the authors for which data was present within at least three studies to answer them. The meta-analysis was conducted based on the proportion of respondents who answered “yes” to the developed questions, as reported in the original studies.

**Table 1:** Questions used to conduct the proportion meta-analysis

| Construct | Question Number | Question |
| --- | --- | --- |
| Knowledge | 1 | Have you received training in family planning counselling? |
|  | 2 | Do you provide family planning counselling to your clients? |
|  | 3 | Have you ever heard of emergency contraception (EC)? |
|  | 4 | The proportion of providers who correctly identify the Copper-intrauterine device as a form of EC. |
|  | 5 | Are you trained to insert intrauterine contraceptive devices (IUCD)? |
|  | 6 | Do you desire more training on IUCD? |
| Attitude and Perceptions | 7 | Do you perceive IUCD to be suitable/safe for HIV-infected women? |
|  | 8 | Do you impose minimum age restrictions for the provision of IUCD? |
|  | 9 | Do you impose minimum parity restrictions for the provision of IUCD? |
|  | 10 | Do you impose minimum age restrictions for the provision of injectable contraceptives? |
|  | 11 | Do you impose minimum parity restrictions for the provision of injectable contraceptives? |

### Quality assessment

The risk of bias for each of the included studies was assessed. Cross-sectional studies were assessed using the ‘instrument for risk of bias in cross-sectional studies of attitudes and practices’(30). The tool allowed for various areas of the study to be assessed such as: how representative the sample was to the general population of interest; the response rate; the impact of missing data; and the reliability of the data instrument used for collection. As one observational study was included, the Newcastle-Ottawa Scale (NOS) was used to assess quality and bias within the study (31). With this tool, the study was assessed based on the selection of study groups and the comparability of the selected groups. Two studies utilising a mixed-method design were also included in the review. These studies were subsequently assessed using the Mixed Methods Appraisal Tool (MMAT) version 18 (32).

### Data synthesis

It was deemed appropriate to combine numerical results from the studies; therefore, a meta-analysis was performed using Stata version 15 (STATA Corp., College Station, TX, USA). Several proportion meta-analyses were conducted based on the questions presented using studies that yielded comparable outcomes. A Stata dataset was created to record the numerators and denominators required to run the ‘metaprop’ command. Where these were not readily available, they were calculated. Heterogeneity was explored using the I^2^-statistic and Chi-squared statistic with results greater than 75% considered to have high heterogeneity (33). For this reason, a random-effects model was used to compensate for the heterogeneity that was seen among the studies (34). For each question, an effect size with 95% CIs were calculated. To further explore the heterogeneity, a sub-group analysis was conducted based on the region in which the study was conducted (East Africa, West Africa, Southern Africa). Publication bias was also explored visually through funnel plots and statistically using Egger’s tests.

## RESULTS

The electronic databases and hand searches yielded a total of 4 448 citations. After removing 832 studies (252 duplicates and 580 published before 2000), 3 616 titles and abstracts were screened. Of these, 3 533 were irrelevant and the remaining 83 full-text articles were assessed for inclusion. Sixty-one were excluded for various reasons (32 reported on an irrelevant outcome not addressing HCW KAP on LARCs, one was a background article, 11 were outside of SSA, one was published before 2000 and 16 did not address LARC methods) and the remaining 22 studies were included in the review. The process is summarised in Figure 1.

**Figure 1.**
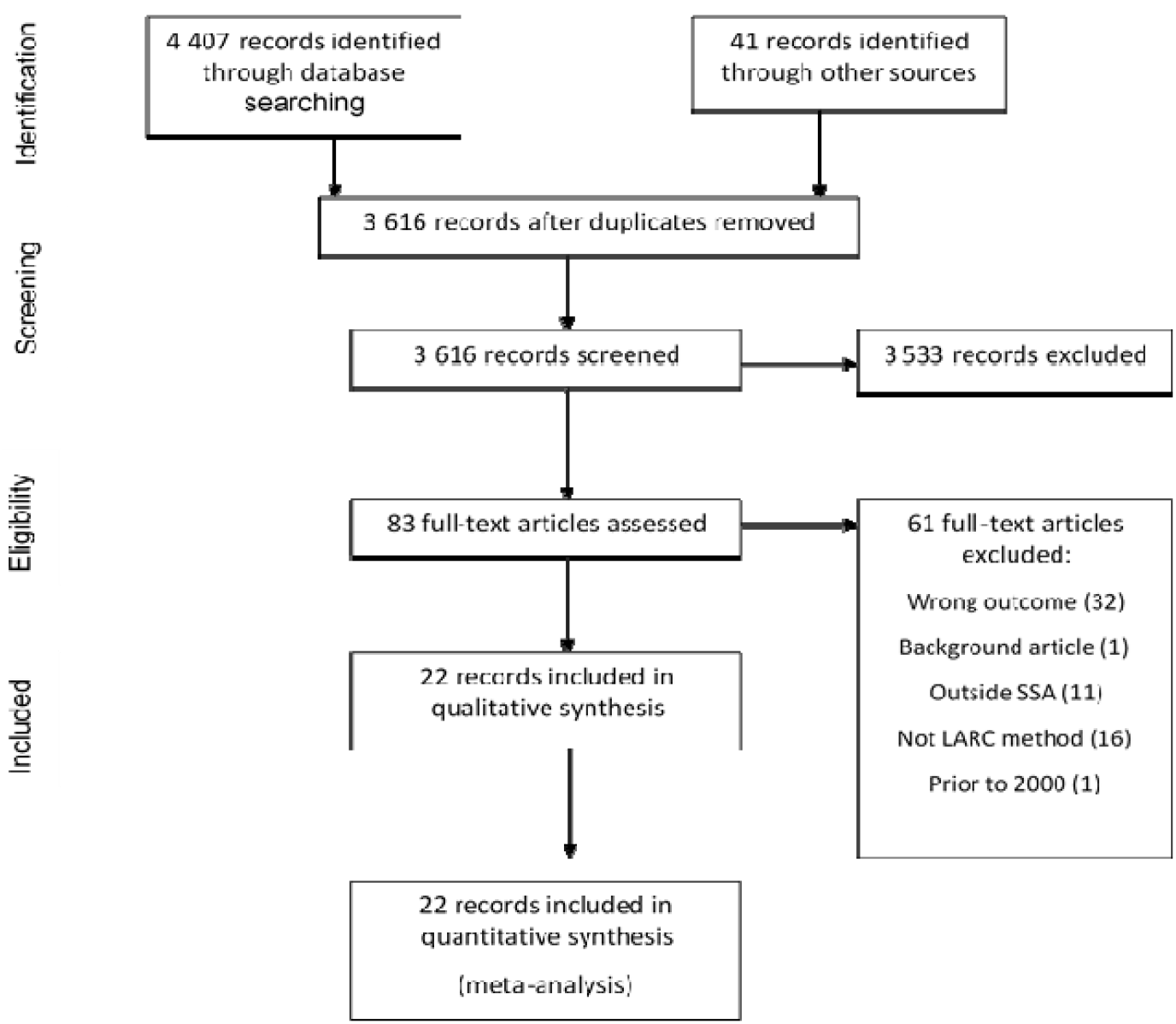
PRISMA flow chart for a systematic review of studies on knowledge, attitudes, and perceptions on long-acting reversible contraceptives in sub-Saharan Africa, 2000-2020.

**Figure 2.**
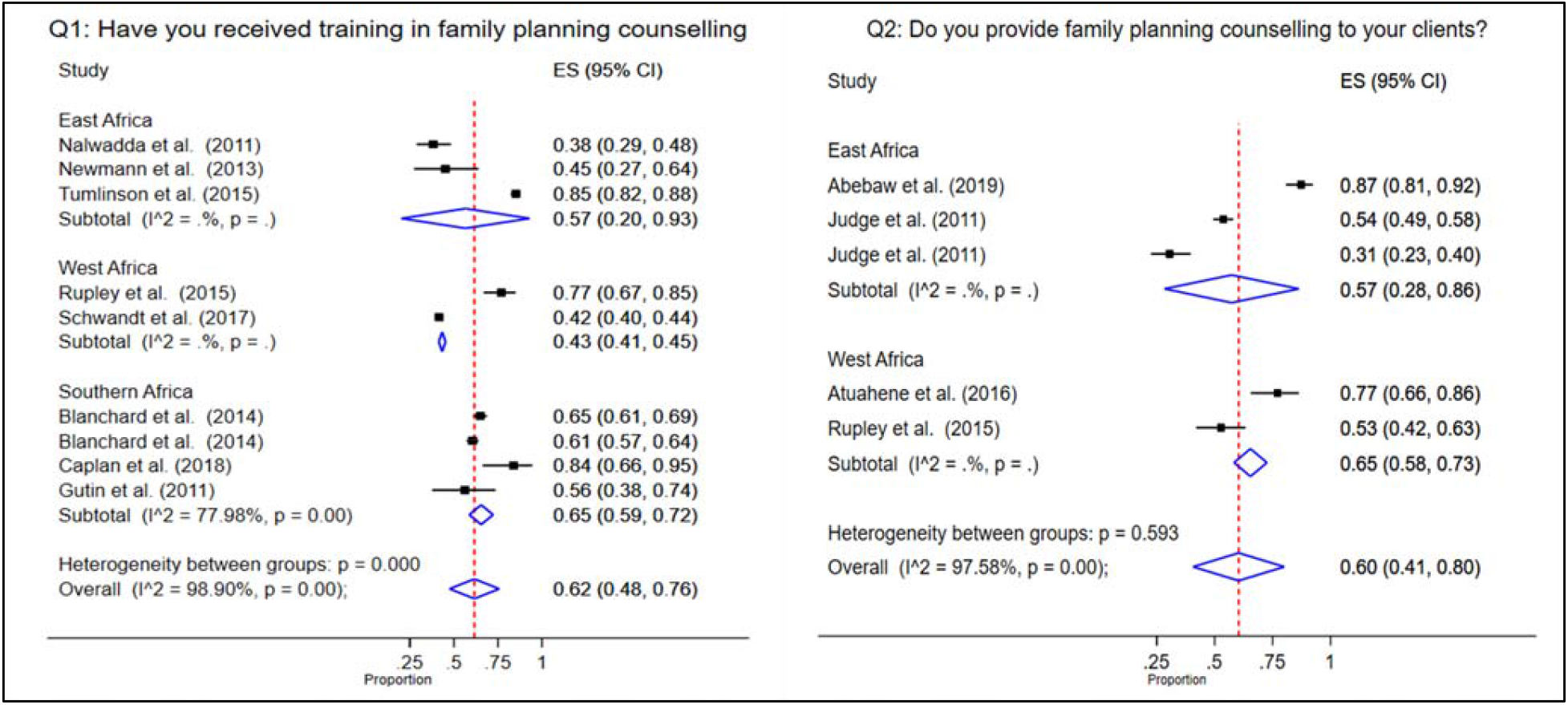
Forest plots showing meta-analyses of studies assessing question 1 and 2 relating to the knowledge of family planning counselling among healthcare workers in sub-Saharan Africa

### Study characteristics

From the 22 included studies published between 2000 and 2019, a total of 11 272 participants were included in this review with the sample size ranging from 31 to 2 377. The studies were conducted across 11 countries including Botswana (35), Ethiopia (36), Ghana (37,38), Kenya (39–41), Malawi (42,43), Nigeria (44–48), Senegal (49), South Africa (50–53), Tanzania (54), Uganda (55,56), and Zimbabwe (50,52). Eleven studies provided information on injectable contraceptives (37,40–43,48–50,54,56,57), 21 on IUCD (36–48,50,51,53–58), and nine on the contraceptive implant (37,41–43,49,50,54,56,58). Additionally, knowledge was reported by 15 studies (36,38,39,41,42,44–47,51–53,55–57), attitudes by 14 (38,39,41–47,51–53,56,57) and perceptions of HCWs by ten studies (37,40–43,49,54,56,57,59,60). A detailed summary of the study characteristics is outlined in Table 2 and Table 3 presents the characteristics of the HCWs included in each study.

**Table 2:**
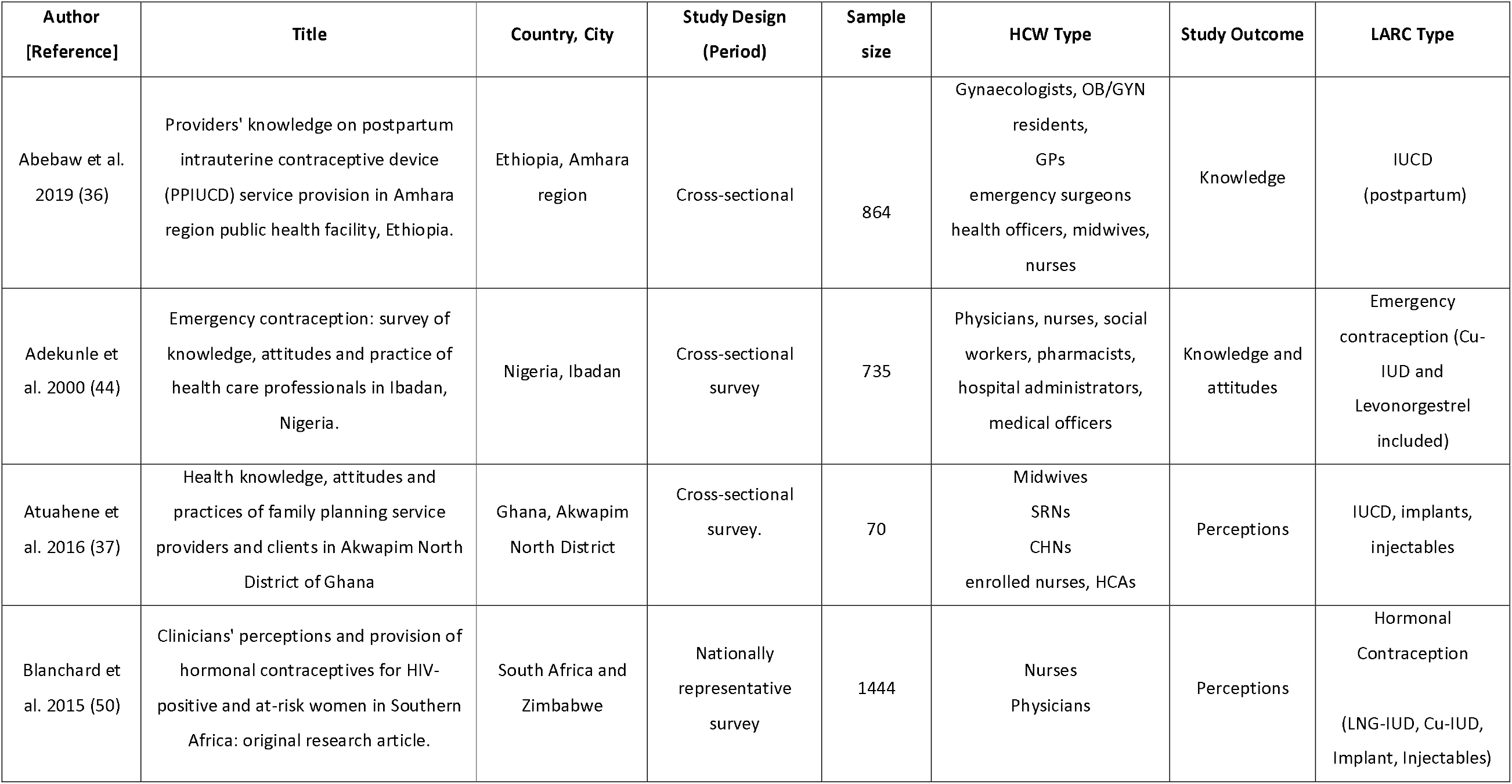
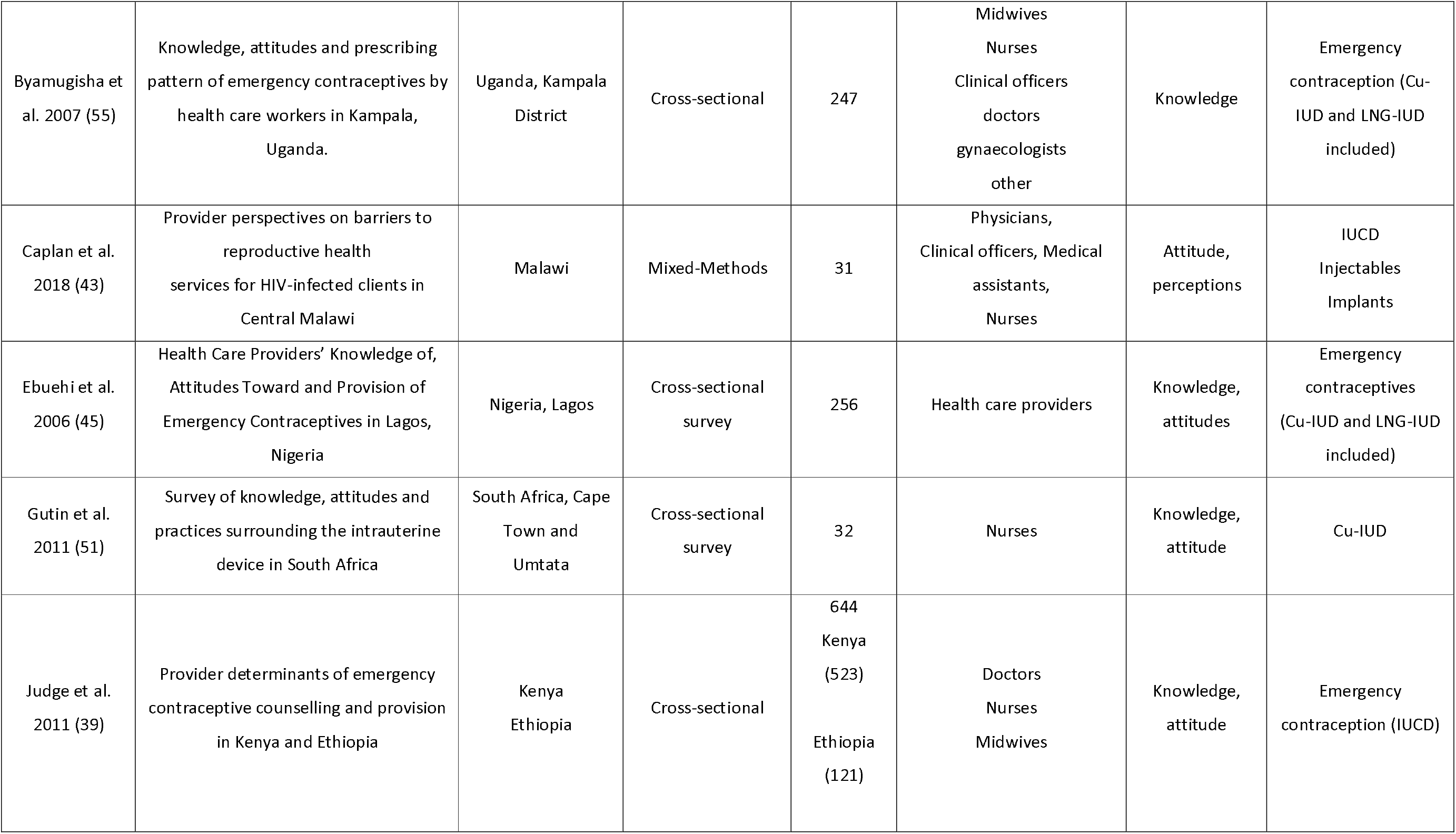
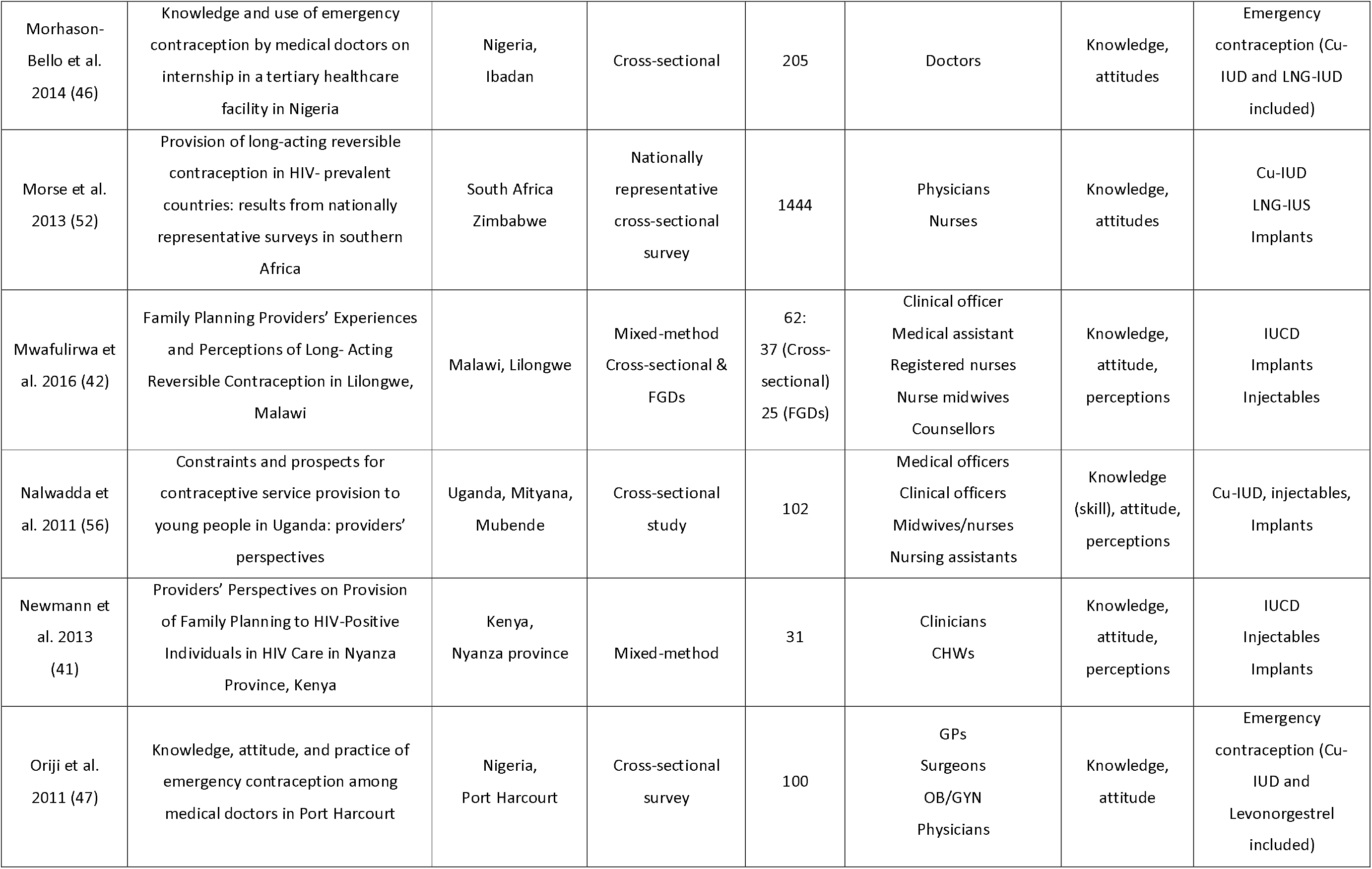
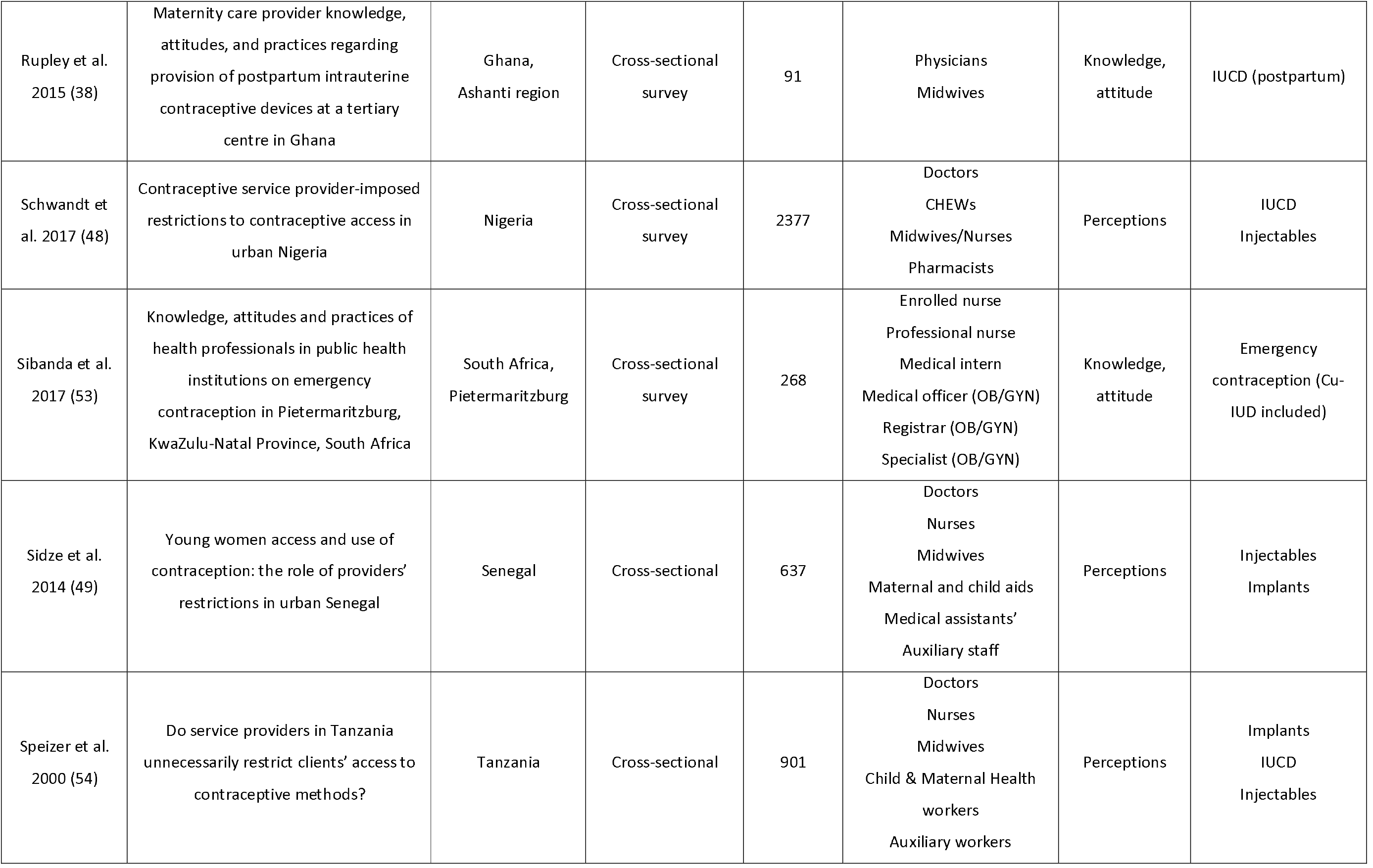
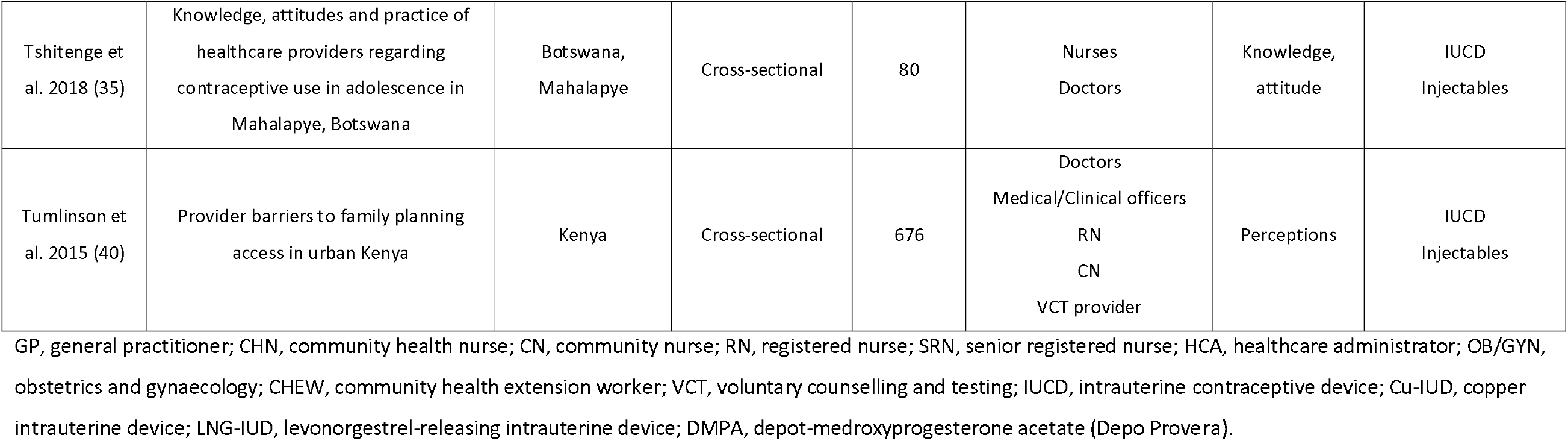
The characteristics of the studies included in the systematic review and meta-analysis of studies assessing the knowledge, attitudes and perceptions of long-acting reversible contraceptives among healthcare workers in sub-Saharan Africa, 2000-2020.

**Table 3:** The characteristics of the healthcare workers included in the studies assessing the knowledge, attitudes, and perception of long-acting reversible contraceptives in sub-Saharan Africa, 2000-2020.

| Author [reference] | Age in years Distribution (n) | Sex | HCW Type Distribution | Religion Distribution | Marital Status Distribution | Experience |
| --- | --- | --- | --- | --- | --- | --- |
| Abebaw et al. 2019 (36) | 18-24 (207)<br>25-34 (570)<br>35-44 (65)<br>45-69 (22) | Male (527)<br>Female (327) | Gynaecologist (5)<br>Resident (10)<br>GP (65)<br>Emergency surgeon (48) health officer (105)<br>midwife (341) nurse (290) | Orthodox (705)<br>Muslim (117) Protestant (21)<br>Other (21) | Married (394)<br>Not married (470) | <5 years (481)<br>5-40 years (299) |
| Adekunle et al. 2000 (44) | <20 (5)<br>21-30 (237) | Male (189)<br>Female (546) | Physicians (183)<br>Nurses (436) | Pentecostal (354) Protestant (124) Muslim (112) Catholic (101) | Single (205) married (485)<br>cohabiting (6) widowed (13) | NR |
|  | 31-40 (313)<br>41-50 (130)<br>51-60 (34)<br>61+ (2)<br>NR* (14) |  | Social workers (50) Pharmacists (38)<br>hospital administrators (16)<br>medical record officers (12). | other (19)<br>None (7)<br>NR (18) | separated (11) divorced (4)<br>NR (11) |  |
| Atuahene et al.<br>2016 (37) | Mean: 31.5 years<br>(SD: 9.8) | Male (13)<br>Female (57) | Midwife (7)<br>SRN (1) CHN (55)<br>enrolled nurse (1)<br>HCA (1), other (3) | NR | NR | <1 year (15)<br>1-2 years (34)<br>3+ years (21) |
| Blanchard et al.<br>2014 (50) | SA: median: 43<br>(29-69)<br>Zimbabwe:<br>median: 39 (20-74) | SA:<br>Male (62),<br>female (547)<br>Zimbabwe:<br>Male (145),<br>female (674) | SA:<br>Nurses (528)<br>Physicians (86)<br>Zimbabwe:<br>Nurses (792)<br>Physicians (38) | NR | NR | SA:<br>FP training<br>(399)<br>HIV training<br>(510)<br>Zimbabwe:<br>FP training<br>(503)<br>HIV training<br>(629) |
| Byamugisha et al.<br>2007 (55) | 21-30 (105)<br>21-30 (84)<br>>40 (33)<br>NR* (25) | NR | Midwives/nurses (185)<br>Clinical officer (16)<br>Doctors (27)<br>Gynaecologist (11)<br>Other (8) | NR | NR | NR |
| Caplan et al. 2018<br>(43) | NR | Female (18)<br>Male (13) | Clinical officer (3)<br>Nurse/ Nurse-technician (20)<br>Medical assistant (8) | Catholic (5)<br>Protestant (20)<br>Other Christian (4) | NR | NR |
|  |  |  |  | Muslim (1) |  |  |
| Ebuehi et al. 2006<br>(45) | NR | Female (132)<br>Male (124) | Physician (116)<br>Nurse (69)<br>Pharmacist (46)<br>CHW (25) | NR |  | Years of<br>experience:<br><10 (37.5%)<br>11-20 (50.4%)<br>21-30 (11.3%)<br>>31 (0.8%) |
| Gutin et al. 2011<br>(51) | NOT REPORTED |  |  |  |  |  |
| Judge et al. 2011<br>(39) | NOT REPORTED |  |  |  |  |  |
| Morhason-Bello<br>et al. 2014 (46) | 20-23 (5)<br>24-27 (127)<br>28-31 (54)<br>32-35 (10)<br>NR (10) | Female (79)<br>Male (126) | Doctors (205) | Christianity (166)<br>Islam (32)<br>Other (4)<br>NR (3) | Single (181)<br>Married (23)<br>Divorced (1) | On internship<br>(205) |
| Morse et al. 2013<br>(52) | Median (range):<br>SA: 43 (23-69)<br>Zimbabwe: 40<br>(20-74) | SA:<br>Female (547)<br>Male (62)<br>Zimbabwe:<br>Female (674)<br>Male (145) | SA:<br>Physicians (86)<br>Nurses (528)<br>Zimbabwe:<br>Physicians (38)<br>Nurses (792) | NR | NR | NR |
| Mwafurirwa et al.<br>2016 (42) | 20-29 (12)<br>30-39 (14)<br>≥40 (11) | Female (24)<br>Male (13) | Clinical officer (1)<br>Medical assistant (4)<br>RN (7)<br>Nurse midwife (23) | NR | NR | NR |
|  |  |  | Counsellor (2)<br>Not reported (25) |  |  |  |
| Nalwadda et al.<br>2011 (56) | NR | NR | Medical officer (6)<br>Clinical officer (6)<br>Midwife/nurse (68)<br>Nursing assistant (22) | NR | NR | FP<br>experience:<br>≤ 5 years (50)<br>6-10 years<br>(32)<br>>10 years (20) |
| Newmann et al.<br>2013<br>(41) | Median (range):<br>33 (30-55) | NR | Clinician (18)<br>CHW (13) | NR | NR | Median<br>(range):<br>3 years (1-26) |
| Orijit et al. 2011<br>(47) | NR | NR | GPs (40)<br>Surgeons (17)<br>Obstetricians/gynaecologist (12)<br>Physicians (9) | NR | NR | 0-5 years (58)<br>6-10 years<br>(30)<br>>10 years (12) |
| Rupley et al. 2015<br>(38) | Median (Range):<br>30 (22-60)<br>**Physicians:** 31<br>(25-51)<br>**Midwives:** 29<br>(22-60) | **Females (65):**<br>Physicians (13)<br>Midwives (52)<br>**Males (26):**<br>Physicians (26)<br>Midwives (26) | Physicians (39)<br>Midwives (52) | NR | NR | NR |
| Schwandt et al.<br>2017 (48) | NR | **Health facility**<br>**Providers:**<br>Female (1313)<br>Male (166)<br>**Pharmacy:**<br>Female (191) | **Health facility Providers (1479):**<br>Doctor (86)<br>CHEW (478)<br>Nurse/Midwife (886)<br>Pharmacist (0)<br>Other (26) | **Health facility providers:**<br>Christian (1019)<br>Islam/Other (460)<br>**Pharmacy:**<br>NR | NR | NR |
|  |  | Male (212)<br>NR (12) | NR (3)<br>**Pharmacy (415):**<br>Doctor (0)<br>CHEW (4)<br>Nurse/Midwife (1)<br>Pharmacist (140)<br>Other (270)<br>NR (3)<br>**PMVs (483):**<br>Doctor (4)<br>Nurse/Midwife (56)<br>CHEW (44)<br>Pharmacists (9)<br>Other (369)<br>NR (1) |  |  |  |
| Sibanda et al.<br>2017 (53) | Mean (range):<br>35.73 (20-61) | Female (228)<br>Male (40) | EN (29)<br>Professional Nurse (172)<br>Medical intern (41)<br>Medical officer (OB/GYN) (11)<br>Registrar (OB/GYN) (14) Specialist<br>(OB/GYN) (1) | NR | NR | NR |
| Sidze et al. 2014<br>(49) | NOT REPORTED |  |  |  |  |  |
| Speizer et al.<br>2000 (54) | NOT REPORTED |  |  |  |  |  |
| Tshitenge et al.<br>2018 (35) | 20-29 (34)<br>30-39 (24) | Female (42)<br>Male (38) | Nurse (50)<br>Doctor (30) | Christian (78)<br>Non-believer/other (2) | Single (40)<br>Married (33) | <5 years (26)<br>5-10 years |
|  | 40-49 (14)<br>50-59 (7)<br>>60 (1) |  |  |  | Living together (6)<br>Widow (1) | (26)<br>11-15 years<br>(12)<br>16-20 years<br>(11)<br>>20 years (5) |
| Tumlinson et al.<br>2015 (40) | 21-29 (218)<br>30-39 (196)<br>40-49 (143)<br>50+ (119) | Female (487)<br>Male (188)<br>NR (1) | Doctors (18)<br>Medical/Clinical officer (115)<br>RN (282)<br>CN (184)<br>VCT provider (77) | Catholic (188)<br>Protestant/Christian (457)<br>Muslim/other (31) | NR | <2 years (71)<br>2-4 years<br>(143)<br>5-9 years<br>(126)<br>10-19 years<br>(135)<br>20+ years<br>(200)<br>NR (1) |

NR, not reported; GP, general practitioner; SRN, senior registered nurse; RN, registered nurse; EN, enrolled nurse; CN, community nurse; CHN, community health nurse; HCA, healthcare administrator; CHW, community health worker; CHEW, community health extension worker; VCT, voluntary counselling and testing; SA, South Africa; FP, family planning; HIV, human immunodeficiency virus.

### Quality of studies

Using the quality assessment tool designed specifically for KAP studies, a total score out of 20 was assigned to each study (30). Eight studies (35–37,45,48,50,52,56) scored at least 15 or higher indicating good quality and eleven (38–40,44,46,47,49,51,53–55) scored 14 or less indicating low quality. The three studies (41–43) that utilised a mixed-method study design were indicative of good quality. The quality of the studies is outlined in Table 4.

**Table 4.** Quality appraisal for the risk of bias among cross-sectional and mixed-method studies assessing the knowledge, attitudes, and perceptions of long-acting reversible contraceptive methods among healthcare workers in sub-Saharan Africa, 2000-2020.

| Risk of Bias for Cross-Sectional Surveys of Knowledge, Attitudes, and Practices |  |  |  |  |  |  |
| --- | --- | --- | --- | --- | --- | --- |
| Study Reference | Is the source population representative of the population of interest? | Is the response rate adequate? | Is there little missing data? | Is the survey clinically sensible? | Is there any evidence for the reliability and validity of the survey instrument? | Total Score (out of 20) |
| Abebaw et al. 2019 (36) | 4 | 4 | 3 | 3 | 3 | 17 |
| Adekunle et al. 2000 (44) | 3 | 4 | 3 | 2 | 1 | 13 |
| Atuahene et al. 2016 (37) | 4 | 4 | 3 | 3 | 2 | 16 |
| Blanchard et al. 2015 (60) | 4 | 3 | 3 | 4 | 4 | 18 |
| Byamugisha et al. 2007 (55) | 4 | 4 | 3 | 3 | 1 | 15 |
| Ebuehi et al. 2006 (45) | 4 | 4 | 3 | 4 | 4 | 19 |
| Gutin et al. 2011 (51) | 3 | 4 | 4 | 1 | 1 | 13 |
| Judge et al. 2011 (39) | 3 | 3 | 2 | 2 | 2 | 12 |
| Morhanson-Bello et al. 2014 (46) | 4 | 4 | 3 | 2 | 1 | 14 |
| Morse et al. 2016 (52) | 4 | 3 | 3 | 4 | 4 | 18 |
| Nalwadda et al. 2011 (56) | 4 | 1 | 4 | 4 | 3 | 16 |
| Orijit et al. 2011 (47) | 3 | 4 | 2 | 1 | 1 | 11 |
| Rupley et al. 2015 (38) | 4 | 2 | 3 | 1 | 3 | 13 |
| Schwandt et al. 2017 (59) | 4 | 3 | 4 | 4 | 3 | 18 |
| Sibanda et al. 2017 (53) | 2 | 4 | 3 | 1 | 3 | 13 |
| Sidze et al. 2014 (49) | 4 | 2 | 2 | 1 | 1 | 10 |
| Speizer et al. 2000 (54) | 3 | 3 | 3 | 2 | 2 | 13 |
| Tshitenge et al. | 4 | 4 | 3 | 4 | 4 | 19 |
| 2018 (57) |  |  |  |  |  |  |
| Tumlinson et al. 2015 (40) | 3 | 4 | 4 | 1 | 2 | **14** |
| **Risk of bias for Mixed-Method studies** |  |  |  |  |  |  |
|  | Is there an adequate rationale for using a mixed-methods design to address the research question? | Are the different components of the study effectively integrated to answer the research question? | Are the outputs of the integration of qualitative and quantitative components adequately interpreted? | Are divergences and inconsistencies between quantitative and qualitative results adequately addressed? | Do the different components of the study adhere to the quality criteria of each tradition of the methods involved? |  |
| Caplan et al. 2018 (43) | No | Yes | Yes | Yes | Yes |  |
| Mwafurirwa et al. 2016 (42) | Yes | Yes | Yes | Yes | Yes |  |
| Newmann et al. 2013 (41) | No | Yes | Yes | Yes | Yes |  |

Key: 4, definitely yes (low risk of bias); 3, probably yes; 2, probably no; 1, definitely yes (high risk of bias).

### Synthesis of results

#### HCW knowledge concerning family planning

Two questions relating to HCW training and HCW willingness to provide family planning counselling to their clients were used to assess their knowledge. The lowest reported proportion (38%) (95% CI: 29%, 48%) of HCWs who had not received family planning training was seen in a study conducted in East Africa (56). The highest proportion of HCWs who had received FP training was 85% (95% CI: 82%, 88%) observed in a Kenyan-based study (40). The overall proportion of HCWs who reported having training in family planning across the nine included studies was 62% (95% CI: 48%, 76%; Chi^2^= 728.06; degrees of freedom [df]= 8; p<0.001; I^2^= 98.9%). Concerning the provision of family planning counselling, the lowest proportion was observed in Ethiopia at only 31% (95% CI: 23%, 40%) in the Addis Ababa, Amhara, Oromiya, Tigray and the Southern Nations, Nationalities and People’s Region’s compared to the highest of 87% (95% CI: 81%, 92%) also in Ethiopia but conducted only in the Amhara Region (36). The analysis, based on five studies, showed that 60% (95% CI: 41%, 80%; Chi^2^= 165.17; df= 4; p<0.001; I^2^= 98.58%) of HCWs provided this service to their clients.

#### Knowledge of emergency contraception

A high proportion (88%; 95% CI: 82%, 94% Chi^2^= 53.93; df= 4; p<0.001; I^2^= 92.58%) of HCWs were aware of EC (from five studies) but only 36% (95% CI: 21%, 51%; Chi^2^= 436.81; df= 7; p<0.001; I^2^= 98.40%) (from eight studies) knew of the Cu-IUDs suitability to be used in clients presenting for EC (Figure 3).

**Figure 3.**
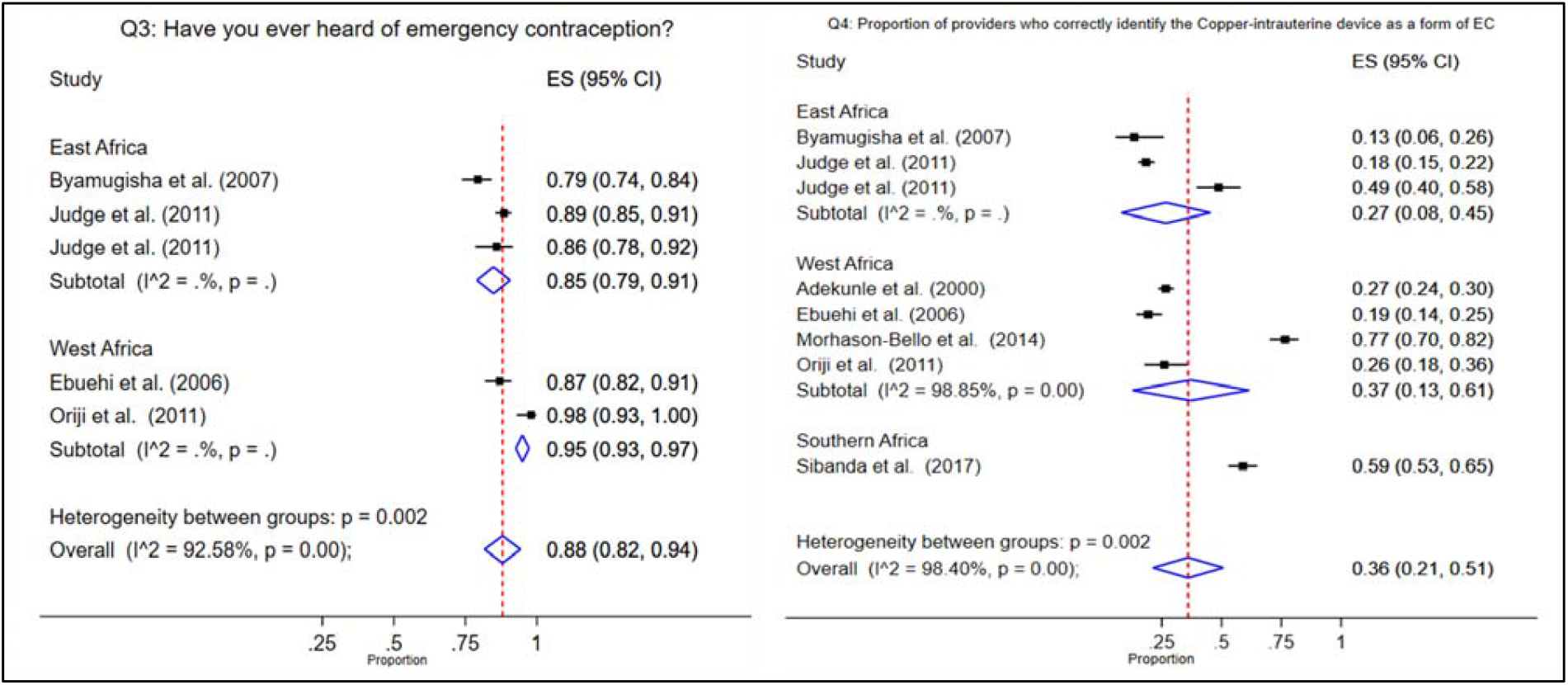
Forest plots showing meta-analyses of studies assessing question 3 and 4 relating to the knowledge of emergency contraception among healthcare workers in sub-Saharan Africa.

**Figure 4.**
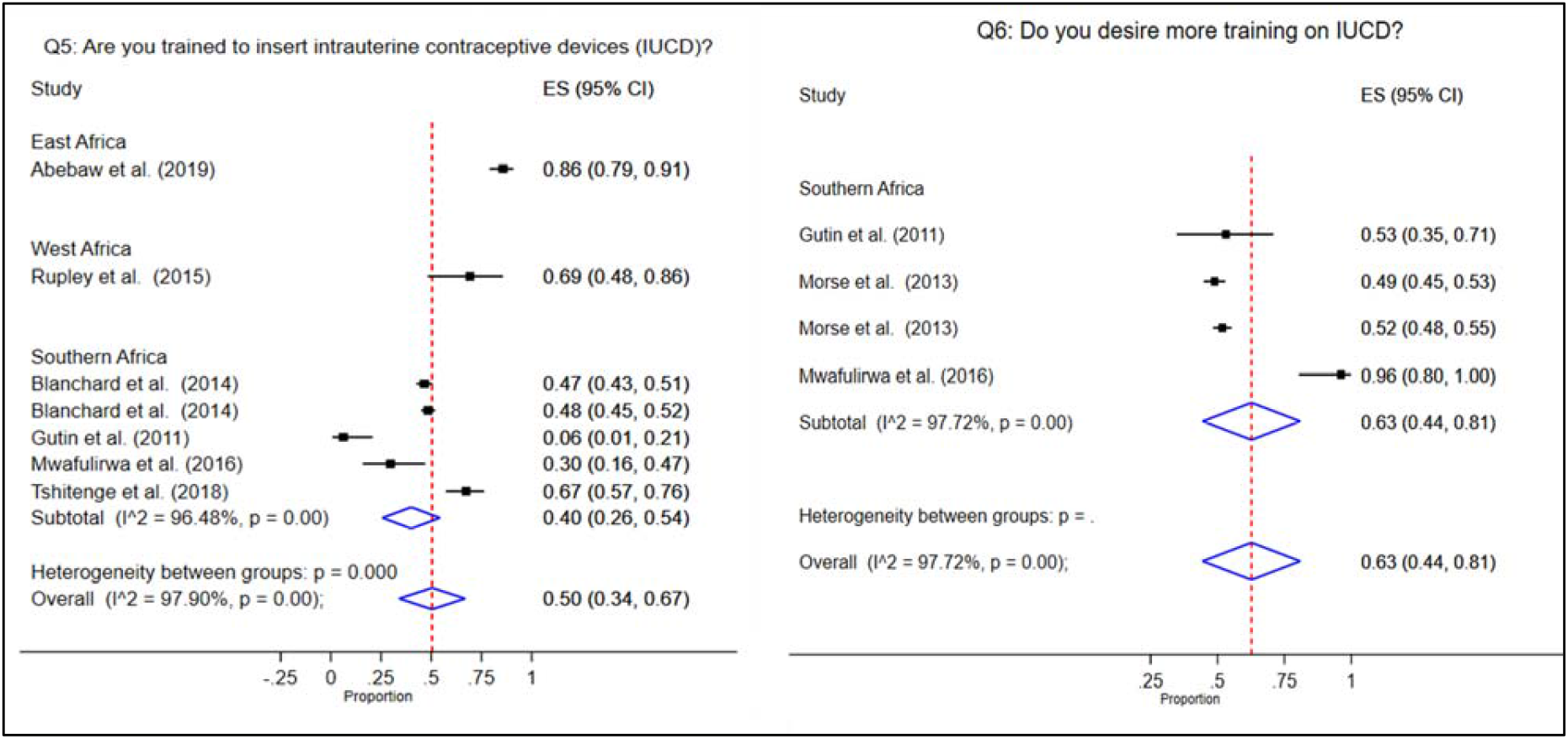

#### Knowledge of IUCD and contraceptive implants

Six studies (35,36,38,42,50,51) reporting on seven different regions were suitable for inclusion in the analysis to investigate the proportion of HCWs trained in IUCD insertion. Studies conducted in South Africa (50, 51), Zimbabwe (50) and Malawi (42) all reported less than 50% of HCW respondents having training on IUCDs. Contrary to this, 86% (95% CI: 79%, 91%) of HCWs in the Amhara Region of Ethiopia were seen to have IUCD insertion training. The overall proportion of HCWs across seven studies in East, West and Southern Africa who had received IUCD insertion training was 50% (95% CI: 34% to 67%; Chi^2^=285.18; df= 6; p<0.001; I^2^= 97.90%). Data from four studies showed that 63% (95% CI: 44%, 81%; Chi^2^=131.63; df= 3; p<0.001; I^2^= 97.72%) of HCWs expressed a desire to receive IUCD insertion training. A meta-analysis on the knowledge relating to the implant was not conducted as there were insufficient studies reporting findings that could be appropriately pooled. However, two studies (42,56) conducted in Malawi and Uganda, respectively, provided some insight into implant knowledge. In Malawi, only 51% of HCWs reported receiving training on implant insertion while 47% of Ugandan HCWs felt competent to provide the method to their clients. Figure 4. Forest plots showing meta-analyses of studies assessing question 5 and 6 relating to the knowledge of intrauterine contraceptive devices among healthcare workers in sub-Saharan Africa.

#### Attitudes and perceptions concerning IUCD and implant contraceptives

The attitudes and perceptions held by HCWs towards IUCD were assessed on their views regarding the safety of the method for HIV-infected women, and their restrictions imposed on its provision based on age and parity. The lowest proportion of HCWs perceiving IUCD as safe for WLWHIV was observed in nationally representative surveys (50) in South Africa (4%; 95% CI: 3%, 6%) and Zimbabwe (5%; 95% CI: 4%, 7%). The highest proportion reported (72%; 95% CI: 53%, 86%) was from HCWs in Cape Town and Umtata, South Africa (51). A meta-analysis of five studies found that 27% (95% CI: 18%, 36%; Chi^2^= 146.64; df= 4; p<0.001; I^2^= 97.27%) of HCWs from the included studies considered IUCD to be suitable and safe for HIV-infected women (Figure 5). With regards to the implant (not included in the meta-analysis as only two studies reported relevant findings), one Zimbabwean study (52) reported 29% of HCWs willing to provide the method to HIV-infected women and 35.5% in a Kenyan study (40).

**Figure 5:**
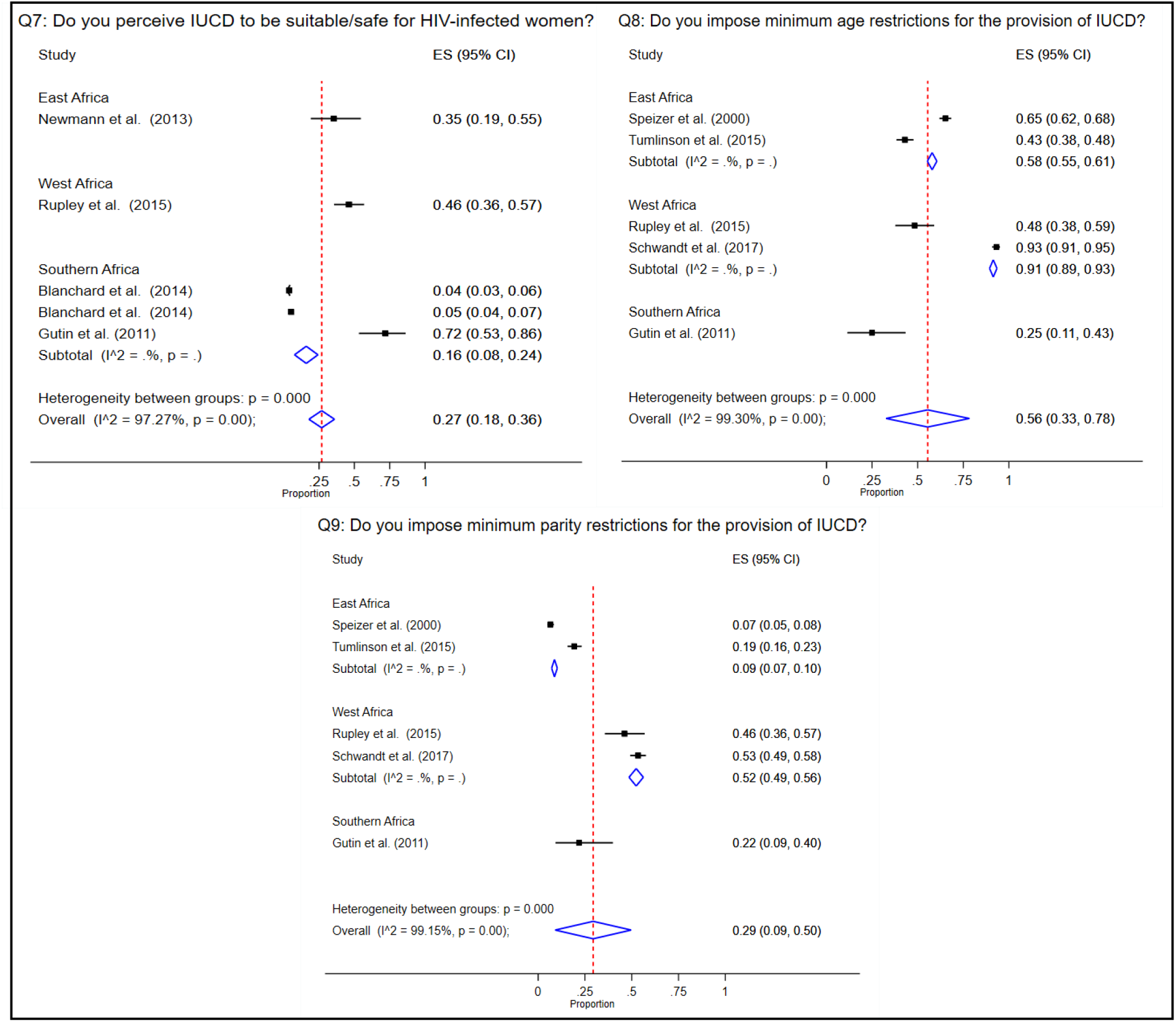
Forest plots showing meta-analyses of studies assessing question 7, 8 and 9 relating to the attitudes and perceptions of intrauterine contraceptive devices among healthcare workers in sub-Saharan Africa.

**Figure 6.**
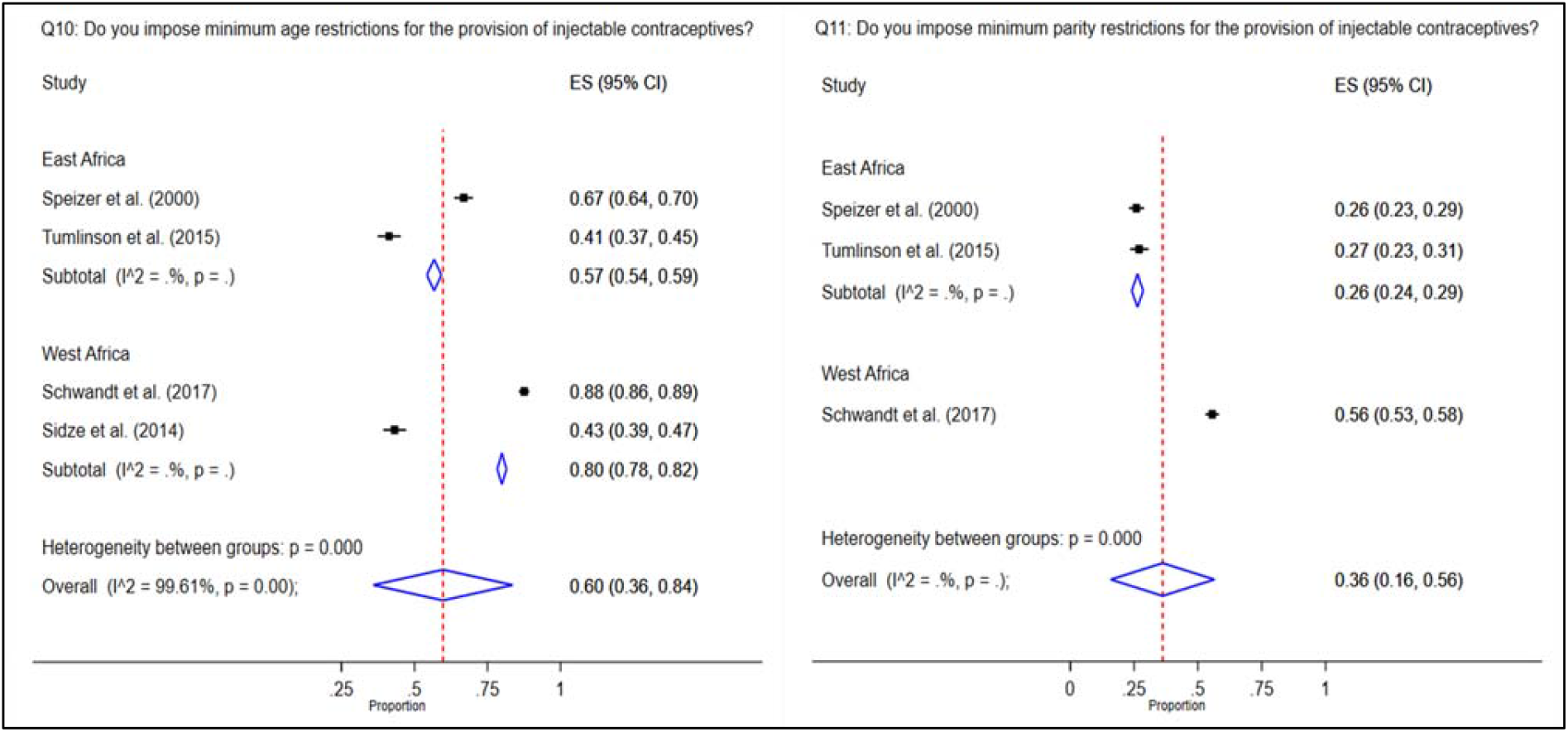
Forest plots showing meta-analyses of studies assessing question 10 and 11 relating to the attitudes and perceptions of injectable contraceptives among healthcare workers in sub-Saharan Africa.

Additional restrictions were observed based on age in which 56% (95% CI: 33%, 78%; Chi^2^= 571.52; df= 4; p<0.001; I^2^= 99.3%) of HCWs (from five studies) imposed minimum age restrictions (most often to adolescent clients) for the provision IUCD. Although not as prevalent, 29% (95% CI: 9%, 50%; Chi^2^= 472.53; df= 4; p<0.001; I^2^= 99.15%) of HCWs (from five studies) suggested that they required a minimum parity before feeling comfortable to provide IUCD to their client. In Senegal (49), 45% of HCWs based in public facilities required a minimum age to provide the implant. Similarly, of 433 Kenyan HCWs, 44% required clients to be at least 20 years of age before inserting an implant (40). Within the same study, of 90 HCWs who reported imposing minimum parity restrictions for the provision of the implant, 56.2% required at least one child, 32.6% required at least two and 11.2% required three or more (40). A meta-analysis for the attitudes and perceptions among HCWs regarding the implant was not conducted due to an insufficient number of studies reporting relevant findings.

#### HCW attitude and perceptions concerning injectable contraceptives

From four studies included in the meta-analysis for question 10, it was observed that 60% (95% CI: 36%, 84%; Chi^2^= 765.55; df= 3; p<0.001; I^2^= 99.61%) of HCWs imposed minimum age restrictions for the provision of injectable contraceptives. Moreover, analysis of three studies showed that 36% (95% CI: 16%, 56%) of HCWs imposed minimum parity restrictions concerning the provision of injectables.

### Publication bias

Through the use of the Egger’s test at a significance level of p< 0.05 and visual exploration of symmetry from funnel plots, it was found that only two questions (7, 10) of the 11 included in the meta-analysis had evidence for publication bias (Figure 7). The results from these two questions should therefore be interpreted with caution.

**Figure 7.**
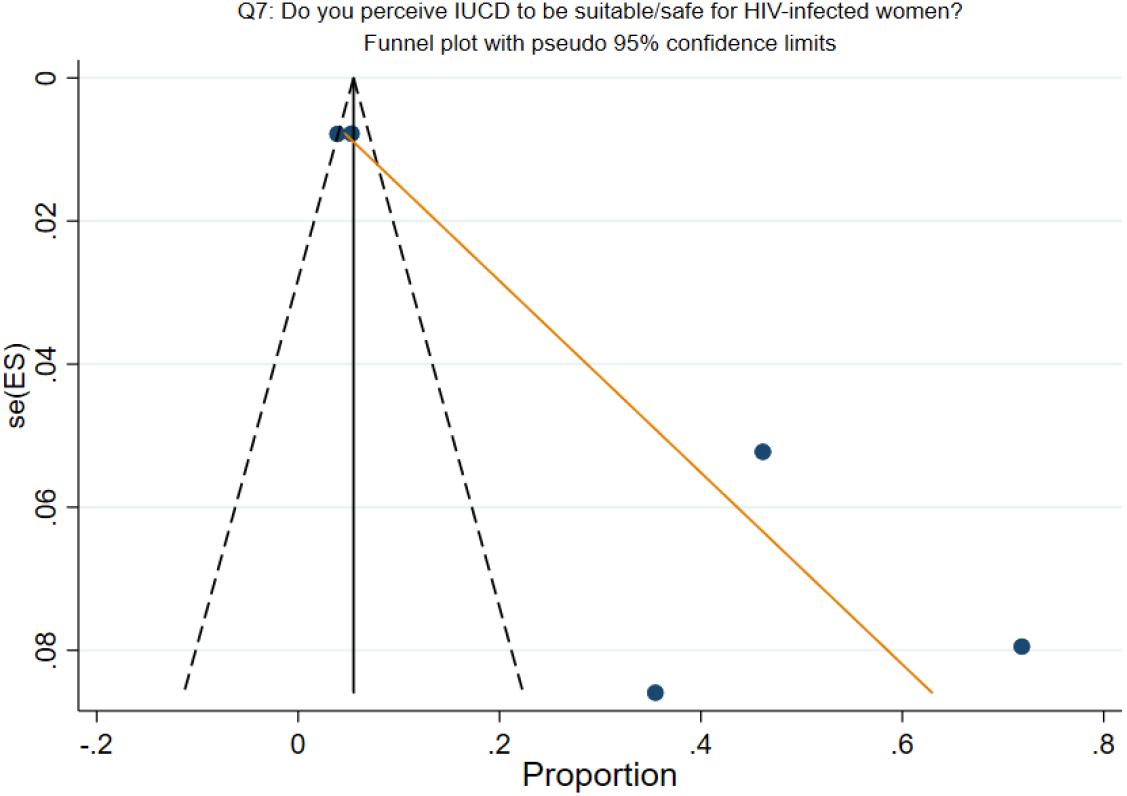
Funnel plot showing the likelihood of publication bias among included studies for question 7 relating to HCWs perceptions of IUCD suitability for HIV-infected women in sub-Saharan Africa.

### Sub-group analysis

A sub-group analysis was conducted based on the region in which the studies were conducted (West, East and, Southern Africa). The analysis showed that the lowest proportion of HCWs who had received FP counselling training was seen in West Africa (43%; 95% CI: 41%, 45%) while 65% (95%CI: 58%, 73%) in the region reported providing it to their clients. In terms of IUCD insertion training, 40% (95% CI: 26%, 54%) had received this in Southern Africa compared to 69% (95% CI: 48%, 86%) in West Africa and 86% (95% CI: 79%, 91%) in East Africa. Further to this, Southern Africa imposed the most restrictions for HIV-infected women with only 16% (95% CI: 8%, 24%) considering the method suitable. However, West Africa imposed the most severe restrictions based on age for IUCD and injectables (91%; 95% CI: 89%, 93%, and 80%; 95% CI: 78%, 82%, respectively). The West African region also had the highest proportion of HCWs who imposed restrictions based on parity at 52% (95% CI: 49%, 56%) for IUCD and 56% (95% CI: 53%, 58%) for injectables.

## DISCUSSION

The slow uptake and underutilisation of LARC methods pose potential barriers to improving SRH across SSA. The burden of unintended pregnancies, in part attributable to the low utilisation of LARC methods, has far-reaching consequences for both maternal and child health outcomes (61). This systematic review and meta-analysis aimed to assess the knowledge, attitudes, and perceptions of LARC methods among HCWs within SSA based on studies published between January 2000 and January 2020. A total of 22 studies were included in the analysis. From the results, knowledge about LARCs was low with 62% of HCWs stating that they readily provide family planning and contraceptive counselling to their clients; this is unsurprising as only 60% of HCWs included in this study had ever received family planning training. This is lower than observed in an Indian study of medical interns and nurses in which 90% and 78%, respectively, had received in-service family planning training (62). However, the findings concur with evidence indicating a high unmet need for quality family planning counselling identified by women across SSA countries, including Ghana (63), Cameroon (64), and Ethiopia(65). The results are concerning as previous research highlights the positive impact that training HCWs in contraceptive counselling has on increasing LARC utilisation (66,67).

A lack of knowledge was also seen in HCWs ability to insert IUCD as only 50% reported having received adequate training to do so. Similarly, a Nigerian study (68) indicated that 56% of HCWs in the study were familiar with IUCDs. However, it is important to note that training in IUCDs is not a guarantee for skill. One Nigerian-based study showed that among HCWs who had received training, many lacked any real-world experience or confidence to insert it (41). The results of this study are not uncommon. A global systematic review of health providers (69) and a systematic review across developed countries (70) both showed that HCWs regularly lacked adequate knowledge of IUCD, particularly on correct candidate selection and insertion ability. There is a clear need to increase HCWs exposure to appropriate IUCD training using conventional medical training programmes as well as real-world training (71). Evidence from multiple studies suggest further and regular training of HCWs (specifically residents, doctors, and nurses) is highly effective in improving knowledge of IUCDs and confidence to provide the method (72–74). This study further indicated that HCWs were prone to imposing unnecessary and medically unsubstantiated restrictions of all LARC methods based on age, parity, and HIV-status. This result is consistent with studies exploring women’s experiences of provider imposed biases when presenting for contraception services (75–77). In this study, only 27% of HCWs considered IUCD safe for HIV-infected women; this is lower than reported in a Nepal-based study showing a proportion of 36% of family planning providers considering IUCD safe for this population (78). With the rates of unintended pregnancies highest among WLWHIV (79) compared to all other groups, there is a need to improve the dialogue between HCWs and their HIV-infected clients desiring effective contraception. Interestingly, HCW KAP toward the female condom, which like LARCs have the benefit of being female-initiated has also been noted to be poor in some settings. For example, among HCWs in Botswana, knowledge of the female condom was poor with only 23% of the 164 respondents reporting being trained on it (80). However, 74% did recommend its use to their clients. Healthcare workers providing care to WLWHIV must be cognisant of their knowledge and attitudes towards both LARCs and barrier methods to prevent unintended pregnancy as well as HIV transmission.

Our study results indicate that adolescents are also negatively affected by HCW attitudes and perceptions. The meta-analysis showed that 56% and 60% of HCWs restricted the provision of IUCDs and injectables based on minimum age, respectively. This is slightly higher than that found in Nepal (78) in which 40% of HCWs restricted the provision of IUCDs to girls under 17 and Pakistan (81) where 50% of physicians restricted access to girls 19 years and younger. The difference in findings may be attributable to the conservative cultural context present within some SSA countries.

Of concern when HCWs refuse LARCs to a particular subset of clients are the resulting behaviour of turning to SARC methods or abandoning contraception altogether. The implication of this is an increase in unintended pregnancies, with the possibility to increase unsafe abortions and ultimately increase the maternal and infant mortality rates. Across SSA, misconceptions and fears among women in communities are known to influence their desire to use LARC methods (82). Our findings, however, show that HCWs knowledge of the methods, attitudes, and perceptions may also have a profound impact on their uptake. With the Family Planning 2020 initiative aiming to increase access to modern contraceptives to an additional 120 million women across the world’s poorest countries, efforts to achieve this should not be hindered, but rather aided by HCWs. Access to reliable and effective contraception underpins a core principle in achieving universal health coverage (UHC); for this reason, a knowledgeable, unbiased health workforce is imperative. Our recommendations to achieve this are grounded in mobilisation to provide quality, routine training for HCWs across all cadres in family planning counselling, LARC insertion skill and education focusing on eliminating misconceptions of appropriate candidates for each method. Secondly, further cadre specific research about the KAP of HCWs is needed as well as more exploration of the KAP among HCWs concerning contraceptive implants, as this is currently limited.

A strength of this study is that, to our knowledge, this is the first systematic review and proportion meta-analysis exploring the KAP of HCWs regarding LARC methods in SSA. Limitations must also be acknowledged. High heterogeneity was observed within each of the analyses. This may be explained by the varying cultural and sociodemographic differences among the HCWs across sub-Saharan Africa. In addition to this, the difference in tools used to assess KAP across different studies and the differences in the way in which they were administered may also have contributed to the high heterogeneity through possible selection bias. Another limitation was the inability to separate the analysis based on the HCW cadre. Lastly, publication bias was evident in question seven and nine of the meta-analysis; these results should, therefore, be interpreted with caution.

## Conclusion

Long-acting reversible contraceptive methods have the potential to greatly increase women’s overall access to contraception, family planning, and birth spacing practices. However, HCWs knowledge of these methods, attitudes and perceptions towards providing them appears to influence women’s choice to utilise them. This can consequently influence the success or failure of strategies that are implemented by governments to increase access to these methods to women. The results from this study have identified a gap in knowledge among HCWs concerning LARCs. There is clear evidence that personal attitudes and perceptions held by HCWs impact their willingness to provide certain methods to a particular set of clients. This was especially seen in the context of HIV-infected women, adolescents, and nulliparous women.

## Data Availability

All data is available upon reasonable request from the corresponding author.

## List of abbreviations

EC: Emergency contraception
FP: Family planning
HCW: Healthcare worker
IUCD: Intrauterine contraceptive device
KAP: Knowledge, attitudes, and perceptions
LARC: Long-acting reversible contraceptive
PRISMA: Preferred reporting items for systematic reviews and meta-analyses
SSA: sub-Saharan Africa

