## Supplemental Search Strategy for "Knowledge, attitudes, and perceptions of long-acting reversible contraceptive (LARC) methods among healthcare workers in sub-Saharan Africa: a systematic review and meta-analysis" (from another version of this preprint: version 1, 2020-10-28)

OVID MEDLINE

1. health worker*.mp. [mp=ti, ab, tx, ct, sh, ot, nm, hw, fx, kf, ox, px, rx, an, ui, sy]

2. general practitioner*.mp. [mp=ti, ab, tx, ct, sh, ot, nm, hw, fx, kf, ox, px, rx, an, ui, sy]

3. primary health worker*.mp. [mp=ti, ab, tx, ct, sh, ot, nm, hw, fx, kf, ox, px, rx, an, ui, sy]

4. nurse*.mp. [mp=ti, ab, tx, ct, sh, ot, nm, hw, fx, kf, ox, px, rx, an, ui, sy]

5. medical practitioner*.mp. [mp=ti, ab, tx, ct, sh, ot, nm, hw, fx, kf, ox, px, rx, an, ui, sy]

6. pharmacist*.mp. [mp=ti, ab, tx, ct, sh, ot, nm, hw, fx, kf, ox, px, rx, an, ui, sy]

7. clinical associate*.mp. [mp=ti, ab, tx, ct, sh, ot, nm, hw, fx, kf, ox, px, rx, an, ui, sy]

8. physician assistant*.mp. [mp=ti, ab, tx, ct, sh, ot, nm, hw, fx, kf, ox, px, rx, an, ui, sy]

9. 1 or 2 or 3 or 4 or 5 or 6 or 7 or 8

10. KAP.mp. [mp=ti, ab, tx, ct, sh, ot, nm, hw, fx, kf, ox, px, rx, an, ui, sy]

11. Knowledge.mp. [mp=ti, ab, tx, ct, sh, ot, nm, hw, fx, kf, ox, px, rx, an, ui, sy]

12. attitude*.mp. [mp=ti, ab, tx, ct, sh, ot, nm, hw, fx, kf, ox, px, rx, an, ui, sy]

13. perception*.mp. [mp=ti, ab, tx, ct, sh, ot, nm, hw, fx, kf, ox, px, rx, an, ui, sy]

14. Practi*.mp. [mp=ti, ab, tx, ct, sh, ot, nm, hw, fx, kf, ox, px, rx, an, ui, sy]

15. availability.mp. [mp=ti, ab, tx, ct, sh, ot, nm, hw, fx, kf, ox, px, rx, an, ui, sy]

16. perspective*.mp. [mp=ti, ab, tx, ct, sh, ot, nm, hw, fx, kf, ox, px, rx, an, ui, sy]

17. training.mp. [mp=ti, ab, tx, ct, sh, ot, nm, hw, fx, kf, ox, px, rx, an, ui, sy]

18. skill.mp. [mp=ti, ab, tx, ct, sh, ot, nm, hw, fx, kf, ox, px, rx, an, ui, sy]

19. 10 or 11 or 12 or 13 or 14 or 15 or 16 or 17 or 18

20. contracep*.mp. [mp=ti, ab, tx, ct, sh, ot, nm, hw, fx, kf, ox, px, rx, an, ui, sy]

21. long acting reversible contracept*.mp. [mp=ti, ab, tx, ct, sh, ot, nm, hw, fx, kf, ox, px, rx, an, ui, sy]

22. copper intrauterine device*.mp. [mp=ti, ab, tx, ct, sh, ot, nm, hw, fx, kf, ox, px, rx, an, ui, sy]

23. IUD.mp. [mp=ti, ab, tx, ct, sh, ot, nm, hw, fx, kf, ox, px, rx, an, ui, sy]

24. contracept* inject*.mp. [mp=ti, ab, tx, ct, sh, ot, nm, hw, fx, kf, ox, px, rx, an, ui, sy]

25. DMPA.mp. [mp=ti, ab, tx, ct, sh, ot, nm, hw, fx, kf, ox, px, rx, an, ui, sy]

26. Medroxyprogesterone acetate.mp. [mp=ti, ab, tx, ct, sh, ot, nm, hw, fx, kf, ox, px, rx, an, ui, sy]

27. Depot Medroxyprogesterone acetate.mp. [mp=ti, ab, tx, ct, sh, ot, nm, hw, fx, kf, ox, px, rx, an, ui, sy]

28. Depo-Provera.mp. [mp=ti, ab, tx, ct, sh, ot, nm, hw, fx, kf, ox, px, rx, an, ui, sy]

29. Progest* contracept*.mp. [mp=ti, ab, tx, ct, sh, ot, nm, hw, fx, kf, ox, px, rx, an, ui, sy]

30. Copper loop.mp. [mp=ti, ab, tx, ct, sh, ot, nm, hw, fx, kf, ox, px, rx, an, ui, sy]

31. Hormonal Birth control.mp. [mp=ti, ab, tx, ct, sh, ot, nm, hw, fx, kf, ox, px, rx, an, ui, sy]

32. ParaGard IUD.mp. [mp=ti, ab, tx, ct, sh, ot, nm, hw, fx, kf, ox, px, rx, an, ui, sy]

33. Norethisterone enanthate.mp. [mp=ti, ab, tx, ct, sh, ot, nm, hw, fx, kf, ox, px, rx, an, ui, sy]

34. NET-EN.mp. [mp=ti, ab, tx, ct, sh, ot, nm, hw, fx, kf, ox, px, rx, an, ui, sy]

35. Nexplanon.mp. [mp=ti, ab, tx, ct, sh, ot, nm, hw, fx, kf, ox, px, rx, an, ui, sy]

36. Implanon.mp. [mp=ti, ab, tx, ct, sh, ot, nm, hw, fx, kf, ox, px, rx, an, ui, sy]

37. Etonogestrel implant.mp. [mp=ti, ab, tx, ct, sh, ot, nm, hw, fx, kf, ox, px, rx, an, ui, sy]

38. Sub-dermal implant.mp. [mp=ti, ab, tx, ct, sh, ot, nm, hw, fx, kf, ox, px, rx, an, ui, sy]

39. rod implant.mp. [mp=ti, ab, tx, ct, sh, ot, nm, hw, fx, kf, ox, px, rx, an, ui, sy]

40. LARC$.mp. [mp=ti, ab, tx, ct, sh, ot, nm, hw, fx, kf, ox, px, rx, an, ui, sy]

41. (Long term and permanent method$).mp. [mp=ti, ab, tx, ct, sh, ot, nm, hw, fx, kf, ox, px, rx, an, ui, sy]

42. 20 or 21 or 22 or 23 or 24 or 25 or 26 or 27 or 28 or 29 or 30 or 31 or 32 or 33 or 34 or 35 or 36 or 37 or 38 or 39 or 40 or 41

43. sub-saharan africa.mp. [mp=ti, ab, tx, ct, sh, ot, nm, hw, fx, kf, ox, px, rx, an, ui, sy]

44. south of the sahara.mp. [mp=ti, ab, tx, ct, sh, ot, nm, hw, fx, kf, ox, px, rx, an, ui, sy]

45. 43 or 44

46. 9 and 19 and 42 and 45

47. remove duplicates from 46

PUBMED

(((((((((((((“health personnel"[MeSH Terms]) OR ("general practitioners"[MeSH Terms]) OR "primary health care worker") OR "nurse") OR "medical practitioner$") OR Pharmacist$) OR "clinical associate$") OR "physician assistant$" OR provider)) AND ((((((((((KAP) OR Knowledge[MeSH Terms]) OR attitude[MeSH Terms]) OR perception[MeSH Terms]) OR practice[MeSH Terms]) OR availability) OR perspective) OR perceptions$) OR training) OR skill) OR Health Knowledge, Attitudes, Practice[MeSH Terms])) AND (((((((((((((((((((((((((((contraception[MeSH Terms]) OR "copper*intrauterine device") OR "intra*uterine device$") OR "intra*uterine coil") OR IUD) OR "Hormonal contraceptive$") OR "contraceptive injectable$") OR "contraceptive devices"[MeSH Terms]) OR "contraceptive agents"[All Fields]) OR "injections"[MeSH Terms]) OR "injectable$"[All Fields]) OR DMPA) OR "Medroxyprogesterone acetate") OR "Depot Medroxyprogesterone acetate") OR "Depo-Provera") OR "Progestin contraceptive$") OR "Copper loop") OR "Birth control") OR "Hormonal Birth control") OR "ParaGard IUD") OR "Norethisterone enanthate") OR "NET-EN") OR "Nexplanon") OR "Implanon") OR "Etonogestrel implant") OR "Sub-dermal implant") OR "rod implant") OR "Long acting reversible contraceptive$") OR "LARC$") OR ("Long term and permanent method$")) AND ((sub-saharan Africa) OR “south of the Sahara”))

SCOPUS

( contraception OR "birth control" OR "family planning" OR contraceptive OR "long acting reversible contraceptives" OR "IUD" OR "copper IUD" OR dmpa OR implanon OR "hormonal contraceptive" OR "hormonal contraceptives" OR "Medroxyprogesterone acetate" OR " Depot Medroxyprogesterone acetate" OR " Depo-Provera" OR " Progestin contraceptive" OR " rod implant" ) AND ( "health workers" OR "health care professionals" OR "health personnel" OR nurses ) AND ( knowledge OR practice OR attitude OR perception OR belief OR skill OR training ) AND ( "sub-saharan africa" OR "south of the sahara" ) AND ( EXCLUDE ( AFFILCOUNTRY , "United States" ) OR EXCLUDE ( AFFILCOUNTRY , "United Kingdom" ) OR EXCLUDE ( AFFILCOUNTRY , "Canada" ) OR EXCLUDE ( AFFILCOUNTRY , "Switzerland" ) OR EXCLUDE ( AFFILCOUNTRY , "Australia" ) OR EXCLUDE ( AFFILCOUNTRY , "Sweden" ) OR EXCLUDE ( AFFILCOUNTRY , "Netherlands" ) OR EXCLUDE ( AFFILCOUNTRY , "India" ) OR EXCLUDE ( AFFILCOUNTRY , "Germany" ) OR EXCLUDE ( AFFILCOUNTRY , "Norway" ) OR EXCLUDE ( AFFILCOUNTRY , "Belgium" ) OR EXCLUDE ( AFFILCOUNTRY , "France" ) OR EXCLUDE ( AFFILCOUNTRY , "Pakistan" ) OR EXCLUDE ( AFFILCOUNTRY , "Brazil" ) OR EXCLUDE ( AFFILCOUNTRY , "Bangladesh" ) OR EXCLUDE ( AFFILCOUNTRY , "China" ) OR EXCLUDE ( AFFILCOUNTRY , "Spain" ) OR EXCLUDE ( AFFILCOUNTRY , "Iran" ) OR EXCLUDE ( AFFILCOUNTRY , "Denmark" ) OR EXCLUDE ( AFFILCOUNTRY , "Thailand" ) OR EXCLUDE ( AFFILCOUNTRY , "Japan" ) OR EXCLUDE ( AFFILCOUNTRY , "Mexico" ) OR EXCLUDE ( AFFILCOUNTRY , "New Zealand" ) OR EXCLUDE ( AFFILCOUNTRY , "Indonesia" ) OR EXCLUDE ( AFFILCOUNTRY , "South Korea" ) OR EXCLUDE ( AFFILCOUNTRY , "Italy" ) OR EXCLUDE ( AFFILCOUNTRY , "Turkey" ) OR EXCLUDE ( AFFILCOUNTRY , "Lebanon" ) OR EXCLUDE ( AFFILCOUNTRY , "Israel" ) OR EXCLUDE ( AFFILCOUNTRY , "Saudi Arabia" ) OR EXCLUDE ( AFFILCOUNTRY , "Ireland" ) OR EXCLUDE ( AFFILCOUNTRY , "Malaysia" ) OR EXCLUDE ( AFFILCOUNTRY , "Viet Nam" ) OR EXCLUDE ( AFFILCOUNTRY , "Austria" ) OR EXCLUDE ( AFFILCOUNTRY , "Chile" ) OR EXCLUDE ( AFFILCOUNTRY , "Papua New Guinea" ) OR EXCLUDE ( AFFILCOUNTRY , "Portugal" ) OR EXCLUDE ( AFFILCOUNTRY , "Russian Federation" ) OR EXCLUDE ( AFFILCOUNTRY , "Singapore" ) OR EXCLUDE ( AFFILCOUNTRY , "Sri Lanka" ) OR EXCLUDE ( AFFILCOUNTRY , "Sudan" ) OR EXCLUDE ( AFFILCOUNTRY , "Taiwan" ) OR EXCLUDE ( AFFILCOUNTRY , "Argentina" ) OR EXCLUDE ( AFFILCOUNTRY , "Cambodia" ) OR EXCLUDE ( AFFILCOUNTRY , "Colombia" ) OR EXCLUDE ( AFFILCOUNTRY , "Haiti" ) OR EXCLUDE ( AFFILCOUNTRY , "Nepal" ) OR EXCLUDE ( AFFILCOUNTRY , "Peru" ) OR EXCLUDE ( AFFILCOUNTRY , "Afghanistan" ) OR EXCLUDE ( AFFILCOUNTRY , "Ecuador" ) OR EXCLUDE ( AFFILCOUNTRY , "Finland" ) OR EXCLUDE ( AFFILCOUNTRY , "Greece" ) OR EXCLUDE ( AFFILCOUNTRY , "Jamaica" ) OR EXCLUDE ( AFFILCOUNTRY , "Luxembourg" ) OR EXCLUDE ( AFFILCOUNTRY , "Nicaragua" ) OR EXCLUDE ( AFFILCOUNTRY , "Oman" ) OR EXCLUDE ( AFFILCOUNTRY , "Dominican Republic" ) OR EXCLUDE ( AFFILCOUNTRY , "Fiji" ) OR EXCLUDE ( AFFILCOUNTRY , "Georgia" ) OR EXCLUDE ( AFFILCOUNTRY , "Guatemala" ) OR EXCLUDE ( AFFILCOUNTRY , "Hong Kong" ) OR EXCLUDE ( AFFILCOUNTRY , "Iraq" ) OR EXCLUDE ( AFFILCOUNTRY , "Laos" ) OR EXCLUDE ( AFFILCOUNTRY , "Myanmar" ) OR EXCLUDE ( AFFILCOUNTRY , "Philippines" ) OR EXCLUDE ( AFFILCOUNTRY , "Poland" ) OR EXCLUDE ( AFFILCOUNTRY , "Qatar" ) OR EXCLUDE ( AFFILCOUNTRY , "Serbia" ) OR EXCLUDE ( AFFILCOUNTRY , "United Arab Emirates" ) OR EXCLUDE ( AFFILCOUNTRY , "Algeria" ) OR EXCLUDE ( AFFILCOUNTRY , "Anguilla" ) OR EXCLUDE ( AFFILCOUNTRY , "Bahamas" ) OR EXCLUDE ( AFFILCOUNTRY , "Bahrain" ) OR EXCLUDE ( AFFILCOUNTRY , "Belize" ) OR EXCLUDE ( AFFILCOUNTRY , "Cape Verde" ) OR EXCLUDE ( AFFILCOUNTRY , "Croatia" ) OR EXCLUDE ( AFFILCOUNTRY , "Cyprus" ) OR EXCLUDE ( AFFILCOUNTRY , "Federated States of Micronesia" ) OR EXCLUDE ( AFFILCOUNTRY , "Iceland" ) OR EXCLUDE ( AFFILCOUNTRY , "Jordan" ) OR EXCLUDE ( AFFILCOUNTRY , "Kazakhstan" ) OR EXCLUDE ( AFFILCOUNTRY , "Kiribati" ) OR EXCLUDE ( AFFILCOUNTRY , "Lithuania" ) OR EXCLUDE ( AFFILCOUNTRY , "Macao" ) OR EXCLUDE ( AFFILCOUNTRY , "Malta" ) OR EXCLUDE ( AFFILCOUNTRY , "Morocco" ) OR EXCLUDE ( AFFILCOUNTRY , "New Caledonia" ) OR EXCLUDE ( AFFILCOUNTRY , "Palestine" ) OR EXCLUDE ( AFFILCOUNTRY , "Panama" ) OR EXCLUDE ( AFFILCOUNTRY , "Slovakia" ) OR EXCLUDE ( AFFILCOUNTRY , "Trinidad and Tobago" ) OR EXCLUDE ( AFFILCOUNTRY , "Ukraine" ) OR EXCLUDE ( AFFILCOUNTRY , "Vanuatu" ) )
